# Antibody responses to SARS-CoV-2 variants LP.8.1, LF.7.1, NB.1.8.1, XFG and BA.3.2 following KP.2 monovalent mRNA vaccination

**DOI:** 10.1101/2025.08.24.25333689

**Authors:** Anass Abbad, Brian Lerman, Jordan Ehrenhaus, Brian Monahan, Gagandeep Singh, Adria Wilson, Stefan Slamanig, Ashley Aracena, Neko Lyttle, Jessica Nardulli, Keith Farrugia, Zain Khalil, Ana Silvia Gonzalez-Reiche, Mia Emilia Sordillo, Weina Sun, Harm van Bakel, Viviana Simon, Florian Krammer

## Abstract

The evolution of severe acute respiratory syndrome coronavirus 2 (SARS-CoV-2) has resulted in antigenically distinct variants that challenge vaccine-induced immunity. The KP.2 monovalent mRNA vaccine was deployed in 2024 to address immune escape by emerging SARS-CoV-2 subvariants. We assessed neutralizing antibody responses in 56 adults with varied exposure histories following KP.2 vaccination against emerging variants including LP.8.1, LF.7.1, NB.1.8.1, XFG, and BA.3.2. While KP.2 vaccination enhanced neutralization against homologous variants, dramatic reductions in neutralizing activity were observed against emerging Omicron variants across all exposure groups. Exposure history showed some influence on neutralization breadth, with self-reported vaccination-only participants exhibiting better cross-neutralization compared to individuals with hybrid immunity. Antigenic cartography revealed substantial antigenic distances between KP.2 and emerging variants, highlighting significant immune escape potential that threatens vaccine protection. Overall, our data suggest that KP.2 boosting predominantly enhances cross-reactive responses imprinted by previously encountered spike antigens, with limited adaptation to antigenically drifted variants.

**IMPORTANCE:** SARS-CoV-2 continues to evolve, producing variants that escape vaccine-induced immunity. The current work shows that KP.2 monovalent vaccination provides limited protection against antigenically distant Omicron variants (LP.8.1, LF.7.1, NB.1.8.1, XFG) and BA.3.2. These findings highlight the ongoing challenge of maintaining vaccine effectiveness against evolving SARS-CoV-2 variants and argue for continuous updating of vaccines.

## OBSERVATION

Severe acute respiratory syndrome coronavirus 2 (SARS-CoV-2) continues evolving in immunologically experienced populations, with emerging variants demonstrating enhanced immune escape. The KP.2 monovalent mRNA vaccine was deployed in 2024 (1).A recombinant protein-based vaccine with the spike of a very similar variant, JN.1, will likely be used in the 2025/2026 season as well even though the mRNA vaccines have been updated to LP.8.1. Understanding neutralizing antibody responses against emerging variants is crucial for informing vaccine strategy and pandemic preparedness. Here, we assessed neutralizing and binding antibody responses in 56 adult study participants with varied SARS-CoV-2 exposure histories following KP.2 monovalent vaccination.

We enrolled 56 healthy adults who received KP.2 monovalent mRNA vaccines and stratified them by exposure profile: vaccination-only (n=16, self-reported, with no strong evidence of any past infection), post-infection boosted (n=11, booster vaccination soon after an infection), and complex hybrid immunity (n=29, ≥2 infections plus ≥3 vaccine doses) (Table S1). Serum samples were collected an average of 29 days post-vaccination. We measured neutralizing antibodies against ancestral WA.1, vaccine-matched KP.2, JN.1 and emerging variants including LP.8.1, LF.7.1, NB.1.8.1, XFG, and BA.3.2 using live and pseudotyped virus microneutralization assays. The tested variants harbor distinct mutation patterns in key antigenic sites (Fig. 1A). BA.3.2, a saltation variant with >50 mutations relative to BA.3, was originally identified in South Africa but has subsequently been detected globally (e.g. Germany, the Netherlands, California, and Australia), reflecting its capacity for transmission despite antigenic divergence (2, 3). FLiRT variants contain critical mutations including R346T, F456L, and Q498R that enhance both ACE2 binding and antibody evasion.

**Fig. 1:**
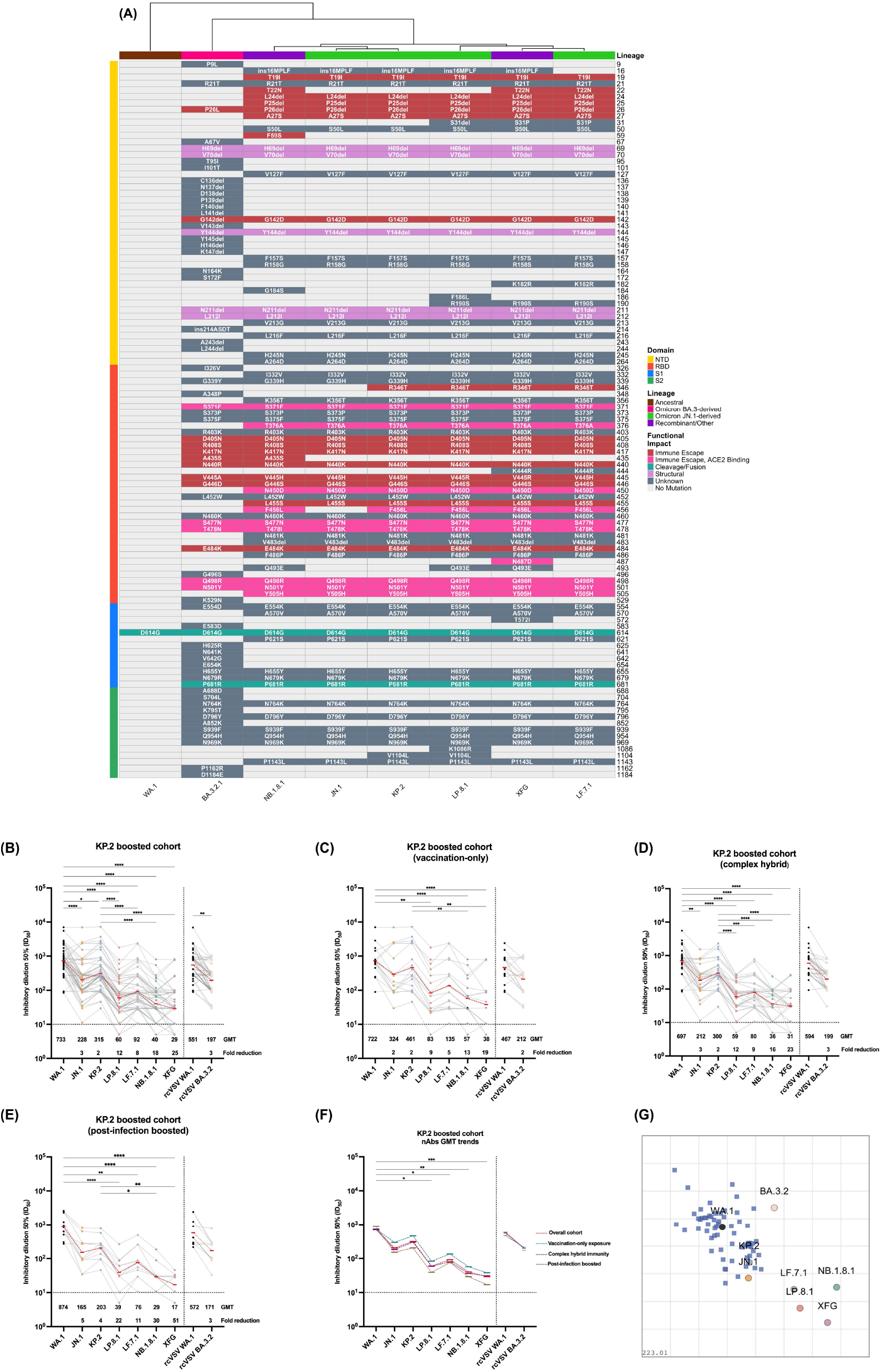
Neutralizing antibody responses and antigenic relationships following KP.2 vaccination. (A) Functional classification of SARS-CoV-2 spike mutations across major variants. Heatmap displaying amino acid mutations in the SARS-CoV-2 spike protein across eight major variants (WA.1, JN.1, KP.2, LP.8.1, LF.7.1, NB.1.8.1, XFG, and BA.3.2) colored by predicted functional impact on receptor binding, antibody escape, and structural stability. Variants are organized by evolutionary lineage, with spike protein domains (N-terminal domain, receptor-binding domain, S1 and S2) indicated. Dendrogram illustrate antigenic similarity relationships based on mutation patterns, with variants clustering by phylogenetic relationship. Color intensity reflects the magnitude of predicted functional consequences for each substitution. **(B) Neutralizing antibody titers following KP.2 monovalent vaccination across exposure groups** against the variant panel for the overall cohort, **(C)** vaccination-only group, **(D)** complex hybrid immunity group and **(E)** recent infection hybrid group. **(F)** Comparative GMT trends across all exposure groups overlaid on a single graph, showing neutralization patterns against each tested variant. Dashed lines connect GMT values for each group. Data are presented as aligned dot plots with individual participant responses connected by lines across the tested virus panel, allowing visualization of individual neutralization patterns. Statistical comparisons between variants within each exposure group are shown above brackets, with significance determined by one-way ANOVA with Dunnett’s multiple comparisons test. (G) **Antigenic cartography reveals spatial relationships between SARS-CoV-2 variants**. Two-dimensional antigenic map constructed from neutralizing antibody titers of the overall KP.2-boosted cohort against the tested variant panel. Each circle represents a virus variant, and each square a unique serum biospecimen, with distances proportional to antigenic differences based on neutralization data. Closely related variants cluster together, while antigenically distinct variants occupy distant positions. Grid lines represent 2-fold changes in neutralizing antibody titers, with each unit corresponding to a 2-fold difference. The map illustrates the antigenic landscape surrounding KP.2, highlighting the substantial antigenic drift of recent omicron variants (LP.8.1, LF.7.1, NB.1.8.1, XFG) and the intermediate positioning of BA.3.2 relative to the vaccine and ancestral strains.

KP.2 vaccination enhanced neutralization against homologous KP.2 (GMT: 315) and closely related JN.1 (GMT: 203) variants (Fig. 1B-E). However, post-KP.2 vaccine sera showed dramatically reduced neutralization against FLiRT variants across all exposure groups. Neutralizing titers against WA.1 (GMT: 733) significantly exceeded those against LP.8.1 (GMT: 60, p<0.0001), LF.7.1 (GMT: 92, p<0.0001), NB.1.8.1 (GMT: 40, p<0.0001), and XFG (GMT: 29, p<0.0001). BA.3.2 showed intermediate neutralization (GMT: 197), representing a 3-fold reduction compared to ancestral strain.

Exposure history influenced neutralization breadth (Fig. 1F). Participants with self-reported vaccination-only immunity exhibited the most potent and broad responses, with highest titers against KP.2 (GMT: 461), JN.1 (GMT: 298), and better neutralization of FLiRT variants compared to other groups (Fig. 1C). Conversely, post-infection boosted participants exhibited the strongest ancestral bias (WA.1 GMT: 874) and lower cross-neutralization: LP.8.1 (GMT: 39), LF.7.1 (GMT: 76), NB.1.8.1 (GMT: 29), and XFG (GMT: 17) (Fig. 1E). Participants with complex hybrid immunity showed intermediate patterns across all variants (Fig. 1D). Importantly, it is unclear if these differences are big enough to be biologically meaningful and they could also be an artifact of a small number of subjects tested. For these reasons, they should not be over-interpreted and require confirmation.

Antigenic cartography quantified these escape patterns (4, 5), revealing that emerging variants occupy distant positions in antigenic space relative to KP.2 (Fig. 1G). FLiRT variants clustered at antigenic distances exceeding three units from KP.2 (representing >8-fold neutralization reductions), while BA.3.2 occupied an intermediate but distinctly separate position. This spatial organization directly correlates with neutralization data and shows substantial immune escape potential that threatens protection.

Our findings reveal significant challenges posed by continued SARS-CoV-2 antigenic evolution. The dramatic reduction in neutralizing activity against FLiRT variants, driven by mutations in critical antigenic sites, highlights the enhanced immune escape capabilities of these variants. The unexpected finding that vaccination-only participants showed better cross-neutralization compared to individuals with hybrid immunity challenges conventional assumptions about hybrid immunity advantages. However, this finding should not be over-interpreted as the differences were small and it is not clear if they represent biologically meaningful differences. It also has to be mentioned that these individuals had no self-reported infections but could of course have had asymptomatic/undetected infections. Furthermore, the finding could also be an artifact due to the small sample size.

Only and intermediate neutralization reduction was observed for BA.3.2, despite its extensive mutation profile. Our finding here is in contrast to another report (3) that show more drastic reduction in neutralization. This difference may be explained by different assay settings. Specifically, we used an assay that assesses multicycle replication in the presence of serum while other reports essentially only looked at initial entry inhibition. The results are in better agreement with a study from Germany (6), even though the variant comparisons are not exactly the same. Our results may explain why this variant has not achieved high transmission rates globally. This could reflect a balance between immune escape and viral fitness costs associated with extensive mutations.

These data highlight the need for adaptive vaccine approaches. Future strategies should consider targeting conserved epitopes or employing alternative delivery methods such as intranasal vaccination to enhance mucosal protection (7). Continuous antigenic surveillance and rapid vaccine updates will be essential as SARS-CoV-2 continues evolving in immunologically experienced populations.

This study has several limitations. The sample size, while adequate for detecting major differences, may limit detection of subtle variations between subgroups. The pseudotype system used for BA.3.2 testing may not fully recapitulate live virus neutralization. Additionally, the durability of these responses beyond the measured timepoint remains unknown. Here, only serological responses were evaluated; cellular immunity, which significantly contributes to protection, was not assessed. Despite these limitations, the observed trends in immune escape and antibody quality remain relevant for informing ongoing vaccine updates and public health strategies.

## Data Availability

Data will be made available upon request to the corresponding author.

## Acknowledgements

This study would not have been feasible without the sustained assistance of the generous research participants, and we thank them for their contribution. We are also thankful to the Mount Sinai Pathogen Surveillance Program for providing representative Omicron viral isolates in a timely manner.

This study was supported by the NIAID Centers of Excellence for Influenza Research and Response (CEIRR, 75N93021C00014, FK and VS), the NIAID Collaborative Influenza Vaccine Innovation Centers (CIVIC, 75N39019C00051, FK and VS) and institutional funding. This project has been funded in part with Federal funds from the National Cancer Institute, National Institutes of Health, under Contract No. 75N91019D00024, Task Order No. 75N91021F00001. The content of this publication does not necessarily reflect the views or policies of the Department of Health and Human Services, nor does mention of trade names, commercial products or organizations imply endorsement by the U.S. Government.

The Conventional Biocontainment Facility (CBF) is a NIHBSL3/BSL3 facility that is part of the BSL-3 Biocontainment CoRE. This Core is supported by subsidies from the ISMMS Dean’s Office and by investigator support through a cost recovery mechanism. Research reported in this publication was supported by the National Institute of Allergy and Infectious Diseases of the National Institutes of Health under Award Number G20AI174733 (R.A. Albrecht). The content is solely the responsibility of the authors and does not necessarily represent the official views of the National Institutes of Health.

## Author Contributions

AA, BL, JE, AW and GS conducted experiments. AA, BL and BM contributed to data analysis. BM and KS assisted with sample processing and data collection. SS and WS shared rcVSV-spike pseudoviruses. FK and VS conceived and supervised the study, designed experiments, analyzed data. AA, VS and FK wrote the manuscript. All authors reviewed and approved the final manuscript.

## fig. legend

**Table S1.** Demographics, SARS-CoV-2 infection and vaccination summary table of post-KP.2 boosted cohort. Summary information for a total of 56 individuals collected serum samples (average of 29 days) following mRNA monovalent KP.2 vaccine booster.

|  | Vaccination Only | Post-Infection Boosted | Complex Hybrid Immunity | Total |
| --- | --- | --- | --- | --- |
| N total | 16 | 11 | 29 | 56 |
| Age | avg (min – max)<br>44.8 (20 – 75) | avg (min – max)<br>40.5 (25 – 62) | avg (min – max)<br>41.4 (23 – 81) | avg (min – max)<br>42.2 (20 – 81) |
| Sex at Birth | n (% group) | n (% group) | n (% group) | n (% total) |
| Female | 11 (68.75%) | 8 (72.73%) | 23 (79.31%) | 42 (75%) |
| Male | 5 (31.25%) | 3 (27.27%) | 6 (20.69%) | 14 (25%) |
| Bivalent Boost | n (% group) | n (% group) | n (% group) | n (% total) |
| Yes | 11 (68.75%) | 10 (90.91%) | 19 (65.52%) | 40 (71.43%) |
| No | 5 (31.25%) | 1 (9.09%) | 10 (34.48%) | 16 (28.57%) |
| XBB Boost | n (% group) | n (% group) | n (% group) | n (% total) |
| Yes | 9 (56.25%) | 6 (54.55%) | 24 (82.76%) | 39 (69.64%) |
| No | 7 (43.75%) | 5 (45.45%) | 5 (17.24%) | 17 (30.36%) |
| Infection | n (% group) | n (% group) | n (% group) | n (% total) |
| Yes | 0 (0%) | 11 (100%) | 29 (100%) | 42 (75%) |
| No | 16 (100%) | 0 (0%) | 0 (0%) | 14 (25%) |
| KP.2 vaccination type | n (% group) | n (% group) | n (% group) | n (% total) |
| Pfizer | 12 (75%) | 4 (36.36%) | 19 (65.52%) | 35 (62.5%) |
| Moderna | 4 (25%) | 7 (63.64%) | 10 (34.48%) | 21 (37.5%) |

## Methods

### Human cohort description

De-identified human serum samples were collected from 56 healthy participants enrolled in the following observational studies: PARIS (Protection Associated with Rapid Immunity to SARS-CoV-2, IRB20-03374) and the observational longitudinal clinical sample collection from patients with emerging viral infections (IRB-17-00791/STUDY-16-01215) at the Icahn School of Medicine at Mount Sinai. All participants received KP.2 monovalent mRNA vaccines and were stratified based on their SARS-CoV-2 exposure history. These studies were approved by the Mount Sinai Hospital Institutional Review Board and all participants provided written informed consent prior to sample and data collection. Information regarding COVID-19 vaccination, SARS-CoV-2 infection were participant reported. Demographics and immune history analysis are shown in Table 1.

### Cell culture

Vero-E6 cells expressing transmembrane protease serine 2 (TMPRSS2) (BPS Biosciences, cat. no. 78081) were maintained in Dulbecco’s modified Eagle’s medium (DMEM; Gibco, cat. no. 11965092) supplemented with 10% heat-inactivated fetal bovine serum (FBS; Gibco, cat. no. A5256801), 1% minimum essential medium with non-essential amino acids (Gibco, cat. no. 11140050), 100 U/mL penicillin and 100 μg/mL streptomycin (Gibco, cat. no. 15140122), 100 μg/mL normocin (InvivoGen, cat. no. ant-nr), and 3 μg/mL puromycin (InvivoGen, cat. no. ant-pr). Baby hamster kidney cells expressing angiotensin converting enzyme 2 (BHK-ACE2) cells were cultured in DMEM supplemented with 10% FBS and 1% penicillin/streptomycin. Expi293F cells (Gibco, cat. no. A14527) were maintained in Expi293 Expression Medium (Gibco, cat. no. A1435102).

### Replication competent SARS-CoV-2 isolates

The SARS-CoV-2 isolate USA-WA.1/2020 was used as a wild-type/ancestral reference (BEI Resources; NR-52281). The following viral isolates from the Omicron lineage were provided by the Mount Sinai Pathogen Surveillance Program: hCoV-19/USA/NY-MSHSPSP-PV96109/2023 (JN.1), hCoV-19/USA/NY-MSHSPSP-PV112116/2024 (KP.2), hCoV-19/USA/NY-MSHSPSP-PV301922/2024 (LP.8.1), hCoV-19/USA/NY-MSHSPSP-PV301501/2025 (LF.7.1), hCoV-19/USA/NY-MSHSPSP-PV304487/2025 (NB.1.8.1), and hCoV-19/USA/NY-MSHSPSP-PV304164/2025 (XFG).

### Replication competent SARS-CoV-2 neutralization assay

Neutralizing antibody titers against SARS-CoV-2 variants (WA.1, JN.1, KP.2, LP.8.1, LF.7.1, NB.1.8.1, and XFG) were measured using a multicycle microneutralization assay in a BSL-3 facility. Vero-E6 TMPRSS2 cells (2×105 cells/well) were seeded in 96-well plates 24 hours prior to infection. Heat-inactivated sera were 3-fold serially diluted starting at 1:10 in modified Eagle’s medium (1xMEM) and incubated with 10,000 50% tissue culture infectious dose (TCID_50_) of each virus for 1 hour at room temperature. Virus-serum mixtures were transferred to cell plates and incubated for 1 hour at 37°C. After removing inoculum, 1xMEM supplemented with 2% FBS was added, and plates were incubated for 48 hours at 37°C. Cells were fixed with 10% formaldehyde overnight at 4°C, permeabilized with 0.1% Triton X-100, and blocked with 3% bovine serum albumin (BSA) in PBS. Biotinylated anti-SARS-CoV nucleoprotein mAb 1C7C7 (1 μg/mL) was added for 2 hour, followed by HRP-conjugated streptavidin (1:2,000) for 1 hour. OPD substrate was added for 10 minutes, stopped with 3 M HCl, and optical density was measured at 490 nm. The 50% inhibitory dilution (ID_50_) was calculated using non-linear regression analysis with 100% and 0% constraints.

### Pseudotyped virus neutralization assay

Neutralizing antibody titers against WA.1 and BA.3.2 variants were measured using replication-competent vesicular stomatitis virus expressing SARS-CoV-2 spike in place of VSV-G (rcVSV-eGFP-CoV2-S) as previously described ^2^. Virus stocks were expended on BHK-ACE2 cells and titrated on Vero-E6 TMPRSS2 cells prior to use. One day before the assay, Vero-E6 TMPRSS2 cells (2×105 cells/well) were seeded in 96-well plates. Heat-inactivated sera were initially diluted 1:10 in 1× minimum essential medium (MEM) followed by 3-fold serial dilutions. Diluted sera were incubated with 10,000 TCID_50_ of each VSV-spike pseudotyped virus under conditions identical to those used for live virus microneutralization assays. Following 48-hour incubation, cells were fixed and permeabilized as described above. Fixed cell plates were immunostained with anti-VSV-N monoclonal antibody (10G4, cat. no. EB0009) at a final dilution of 1:3,000 for two hours at room temperature, followed by HRP-conjugated anti-mouse secondary antibody (Rockland, cat. no. 610-1102) at a final dilution of 1:3,000 for 1 hour. OPD substrate was added for 10 minutes, the reaction was stopped with 3 M HCl, and optical density was measured at 490 nm. The ID_50_ titers were calculated as described above.

### Antigenic cartography

Antigenic maps were constructed from neutralization data using established multidimensional scaling methods as previously described ^3,4^. Neutralizing antibody titers were converted to antigenic distances, where each unit corresponds to a 2-fold difference in neutralization capacity. Variants and sera were positioned in multidimensional space such that Euclidean distances between points accurately reflect antigenic relationships. Maps were optimized to minimize stress between observed and predicted neutralization titers, then projected into two dimensions for visualization. Antigenic cartography maps were generated in R (version 4.3.3) using the Racmacs package (version 1.2.9) ^3^. The number of optimization runs was set to 3,000, and the minimum column basis was set to none to allow for maximum flexibility in map construction. The limit of detection (LoD) for neutralizing antibody titers was established at 1:10, and all titers below this threshold were assigned as “<10” in the titer table for subsequent analysis.

### Statistical analysis

Statistical analyses were performed using Prism 10 software (GraphPad). Log-transformed neutralization titers were compared between groups. Normality was assessed using Shapiro-Wilk tests. For non-normally distributed data, Kruskal-Wallis test with Dunn’s multiple comparisons test was applied. Wilcoxon matched-pairs signed rank test was used for paired comparisons. Geometric mean titers and geometric mean ratios with 95% confidence intervals were calculated for all analyses. Statistical significance was defined as p < 0.05.

